# Illness duration and symptom profile in a large cohort of symptomatic UK school-aged children tested for SARS-CoV-2

**DOI:** 10.1101/2021.05.05.21256649

**Authors:** Erika Molteni, Carole H. Sudre, Liane S. Canas, Sunil S. Bhopal, Robert C. Hughes, Michela Antonelli, Benjamin Murray, Kerstin Kläser, Eric Kerfoot, Liyuan Chen, Jie Deng, Christina Hu, Somesh Selvachandran, Kenneth Read, Joan Capdevila Pujol, Alexander Hammers, Tim D. Spector, Sebastien Ourselin, Claire J. Steves, Marc Modat, Michael Absoud, Emma L. Duncan

## Abstract

**Background:** In children, SARS-CoV-2 is usually asymptomatic or causes a mild illness of short duration. Persistent illness has been reported; however, its prevalence and characteristics are unclear. We aimed to determine illness duration and characteristics in symptomatic UK school-aged children tested for SARS-CoV-2 using data from the COVID Symptom Study, the largest UK citizen participatory epidemiological study to date.

**Methods:** Data from 258,790 children aged 5-17 years were reported by an adult proxy between 24 March 2020 and 22 February 2021. Illness duration and symptom profiles were analysed for all children testing positive for SARS-CoV-2 for whom illness duration could be determined, considered overall and within younger (5-11 years) and older (12-17 years) groups. Data from symptomatic children testing negative for SARS-CoV-2, matched 1:1 for age, gender, and week of testing, were also assessed.

**Findings:** 1,734 children (588 younger, 1,146 older children) had a positive SARS-CoV-2 test result and calculable illness duration within the study time frame. The commonest symptoms were headache (62.2%) and fatigue (55.0%). Median illness duration was six days (vs. three days in children testing negative), and was positively associated with age (*r*_*s*_ 0.19, p<1.e-4) with median duration of seven days in older vs. five days in younger children.

Seventy-seven (4.4%) children had illness duration ≥28 days (LC28), more commonly experienced by older vs. younger children (59 (5.1%) vs. 18 (3.1%), p=0.046). The commonest symptoms experienced by these children were fatigue (84%), headache (80%) and anosmia (80%); however, by day 28 the symptom burden was low (median, two). Only 25 (1.8%) of 1,379 children experienced symptoms for ≥56 days. Few children (15 children, 0.9%) in the negatively-tested cohort experienced prolonged symptom duration; however, these children experienced greater symptom burden (both throughout their illness and at day 28) than children positive for SARS-CoV-2.

**Interpretation:** Some children with COVID-19 experience prolonged illness duration. Reassuringly, symptom burden in these children did not increase with time, and most recovered by day 56. Some children who tested negative for SARS-CoV-2 also had persistent and burdensome illness. A holistic approach for all children with persistent illness during the pandemic is appropriate.

**Research in context:** *Evidence before this study:* SARS-CoV-2 in children is usually asymptomatic or manifests as a mild illness of short duration. Concerns have been raised regarding prolonged illness in children, with no clear resolution of symptoms several weeks after onset, as is observed in some adults. How common this might be in children, the clinical features of such prolonged illness in children, and how it might compare with illnesses from other respiratory viruses (and with general population prevalence of these symptoms) is unclear.

*Added value of this study:* We provide systematic description of COVID-19 in UK school-aged children. Our data, collected in a digital surveillance platform through one of the largest UK citizen science initiatives, show that long illness duration after SARS-CoV-2 infection in school-aged children does occur, but is uncommon, with only a small proportion of children experiencing illness duration beyond four weeks; and the symptom burden in these children usually decreases over time. Almost all children have symptom resolution by eight weeks, providing reassurance about long-term outcomes. Additionally, symptom burden in children with long COVID was not greater than symptom burden in children with long illnesses due to causes other than SARS-CoV-2 infection.

*Implications of all the available evidence:* Our data confirm that COVID-19 in UK school-aged children is usually of short duration and of low symptom burden. Some children do experience longer illness duration, validating their experience; however, most of these children usually recover with time. Our findings highlight that appropriate resources will be necessary for any child with prolonged illness, whether due to COVID-19 or other illness. Our study provides timely and critical data to inform discussions around the impact and implications of the pandemic on paediatric healthcare resource allocation.

## Introduction

To date, severe acute respiratory syndrome-related coronavirus 2 (SARS-CoV-2) has caused >120 million cases of Coronavirus Disease 2019 (COVID-19) and 2.5 million deaths globally.^1^ In adults, SARS-CoV-2 causes a predominantly respiratory illness^2^ of median duration 11 days.^3^ In contrast, children with SARS-CoV-2 infection are often asymptomatic (43%-68%^4^) or have relatively mild symptoms, most commonly cough and fever;^4,5^ and life-threatening illness or death is rare. 1,575 children (aged 0-17 years) hospitalised in England from 19 March 2020 to 3 March 2021 tested positive for SARS-CoV-2 (though COVID-19 may not necessarily have been the reason for hospitalisation^6^) and 29 deaths due to COVID-19 were reported in children and young people across the UK from 1 March 2020 to 29 January 2021 (0.59% of estimated all-cause deaths in this age group during this period^7^).

The pandemic has also seen a new rare condition, multisystem inflammatory syndrome in children (MIS-C), typically presenting 2-4 weeks after acute SARS-CoV-2 infection.^8^ Some adults with SARS-CoV-2 infection experience prolonged illness duration (“Long COVID”).^9,10^ The King’s College London (KCL) COVID Symptom Study (CSS),^11^ involving >4.5 million UK participants, showed that 13.3% adults with COVID-19 had symptoms for ≥4 weeks (LC28) and 4.5% ≥8 weeks (LC56).^3^ Whether some children may also experience prolonged illness duration after COVID-19^10^ and, if so, how this compares with other illnesses, is unclear.

In September 2020, coinciding with UK schools re-opening, governance for CSS data usage was extended to allow analysis of data from children (i.e., individuals aged <18 years). The UK subsequently experienced further waves of the pandemic, during which time there was widespread testing availability for individuals experiencing fever, cough and/or anosmia, in contrast to the limited access during the first wave when testing was mostly restricted to individuals presenting to hospital.^12,13^ Moreover, stay-at-home directives and school closures over the 2020-21 UK winter resulted in unusually low circulation of viruses such as influenza, adenovirus, and respiratory syncytial virus.^14^ However, symptom overlap meant many individuals (adults and children) with respiratory illnesses other than COVID-19 were tested for SARS-CoV-2.

Here we report illness duration, individual symptom prevalence and duration, and symptom burden in UK school-aged children testing positive for SARS-CoV-2, and provide similar data for symptomatic children testing negative during the same period. We also present prevalence and characteristics of long COVID in children.

## Methods

Data were acquired from CSS, through a mobile application launched jointly by Zoe Global Ltd. and KCL on 24 March 2020.^11^ Briefly, individuals report through a smartphone application their symptomatology, any SARS-CoV-2 testing, vaccination, and health care access. Symptom assessment includes direct questions about specific symptoms (Supplementary Table 1) and free-text entry.^11^ Adult contributors can also proxy-report for other persons (children, elderly relatives, etc.). The relationship between contributor and proxy-reported individual(s) is not solicited and there is no data linkage between them. Children aged 16-17 years can use the app independently or be proxy-reported by an adult.

UK data from school-aged children (aged 5-17 years) were available from app launch to 22 February 2021, which latter date corresponds to eight weeks after peak UK SARS-CoV-2 positive specimen date.^15^ The cohort was analysed overall and within two age groups: younger children, aged 5-11 years (UK primary school-aged children) and older children, aged 12-17 years (UK secondary school-aged children).

Aligning with our previous publications in adults,^3,11^ children were considered symptomatic of COVID-19 if proxy-reported with relevant symptoms^11^ (Supplementary Table),^16^ with first symptom(s) presenting within a timeframe of one week before and two weeks after infection confirmation (either SARS-CoV-2 polymerase chain reaction or lateral flow test). Illness duration was calculated from first symptom(s) (having been previously asymptomatic) until recovery (return to asymptomatic state or, if proxy-reporting ceased prior to an asymptomatic report, the final proxy-report). Individuals reported as asymptomatic but subsequently re-reported with symptoms within one week of last symptomatic report were considered still unwell from initial presentation (i.e., having relapsing/remitting illness); calculation of illness duration included these short asymptomatic periods. Children with reporting gaps longer than one week between symptomatic reports were excluded. Individual symptom prevalence and duration were assessed, with individual symptom duration calculated as time between first and last report for that symptom. Symptom burden was calculated as the number of different symptoms reported at least once over a defined timeframe (during first week, first 28 days, ≥28 days until end of illness, and over entire illness duration). Consistent with our previous adult study,^3^ we termed illness with symptoms ≥28 days as LC28; and ≥56 days, LC56. Thus, by virtue of census dates LC28 could only be determined in proxy-reported children whose symptoms commenced on or before 24 January 2021, and for LC56 on or before 29 December 2020 (the peak positive specimen date^15^). Symptom profiles were also assessed in children presenting for hospital-based care (presenting to the emergency department or admitted to hospital), where hospital presentation followed symptom commencement.

Several direct symptom questions were added to the app on 4 November 2020 (Supplementary Table 2). Data from these additional questions are presented in Supplementary Figure 1 but were not included in illness duration or symptom burden calculations for the main analysis.

Free-text reporting was also possible across the entire period. Free-text data were divided into themes using frequency of descriptive words, with each item within the themes independently scrutinised by two clinicians (MAb, ELD) to ensure appropriate categorisation. Individuals reporting free-text symptoms within those themes were then counted. Free-text data are reported here as descriptive statistics (Supplementary Table 3) and were not included in calculating illness duration or symptom burden. Free-text data were also searched for specific neurological terms (e.g., weakness, difficulties with balance, paralysis, seizures, fits, convulsions, paroxysms, tics) and symptoms potentially affecting attention, behaviour, learning, and/or school performance (e.g., anxiety, low mood, and irritability). Symptoms already assessed by direct question (Supplementary Table 1) were excluded from free-text searching, to avoid duplication.

Symptom profile and duration were also assessed in children testing negative for SARS-CoV-2 using a randomly selected control cohort (matched 1:1 for age, gender, and week of testing), and compared to children testing positive, analysed identically.

Proxy-reporting density was defined as the number of episodes of proxy-reporting over the illness duration, and persistence as proxy-reporting until return to healthy state.

Prevalence data for common winter circulating viruses were obtained from the Public Health England weekly national influenza and COVID-19 surveillance report.^14^

Data are presented using descriptive statistics. Results are presented as median and interquartile ranges. Due to rarity (with most percentages <5%) confidence intervals were calculated using Poisson distribution. Comparisons of data between groups used Wilcoxon signed-rank test or Chi-squared/Fisher’s exact tests, as appropriate. Spearman correlation testing was used to assess correlation of illness duration with age.

## Results

Recruitment is outlined in Figure 1. Overall, 258,790 children aged 5-17 years were proxy-reported between 24 March 2020 and 22 February 2021. A positive SARS-CoV-2 test result was reported in 6,975 children, of whom 1,912 (666 younger, 1,246 older) children had a calculable illness duration and requisite proxy-report logging. As only 36 of these children had illness onset prior to 1 September 2020 (which date corresponded to return-to-school), and with limited community access to testing early in the UK pandemic,^13^ analyses were restricted to children with illness onset after 1 September 2020. Illness onset had to commence on or before 24 January 2021 for illness duration ≥4 weeks to be evident, per LC28 definition;^3^ 1,734 (588 younger, 1,146 older) children were proxy-logged within the requisite timeframe. Similarly, 1,379 (445 younger; 934 older) children had symptoms commencing on or before 29 December 2020, allowing illness duration ≥8 weeks to be evident, per LC56 definition.^3^

**Figure 1.**
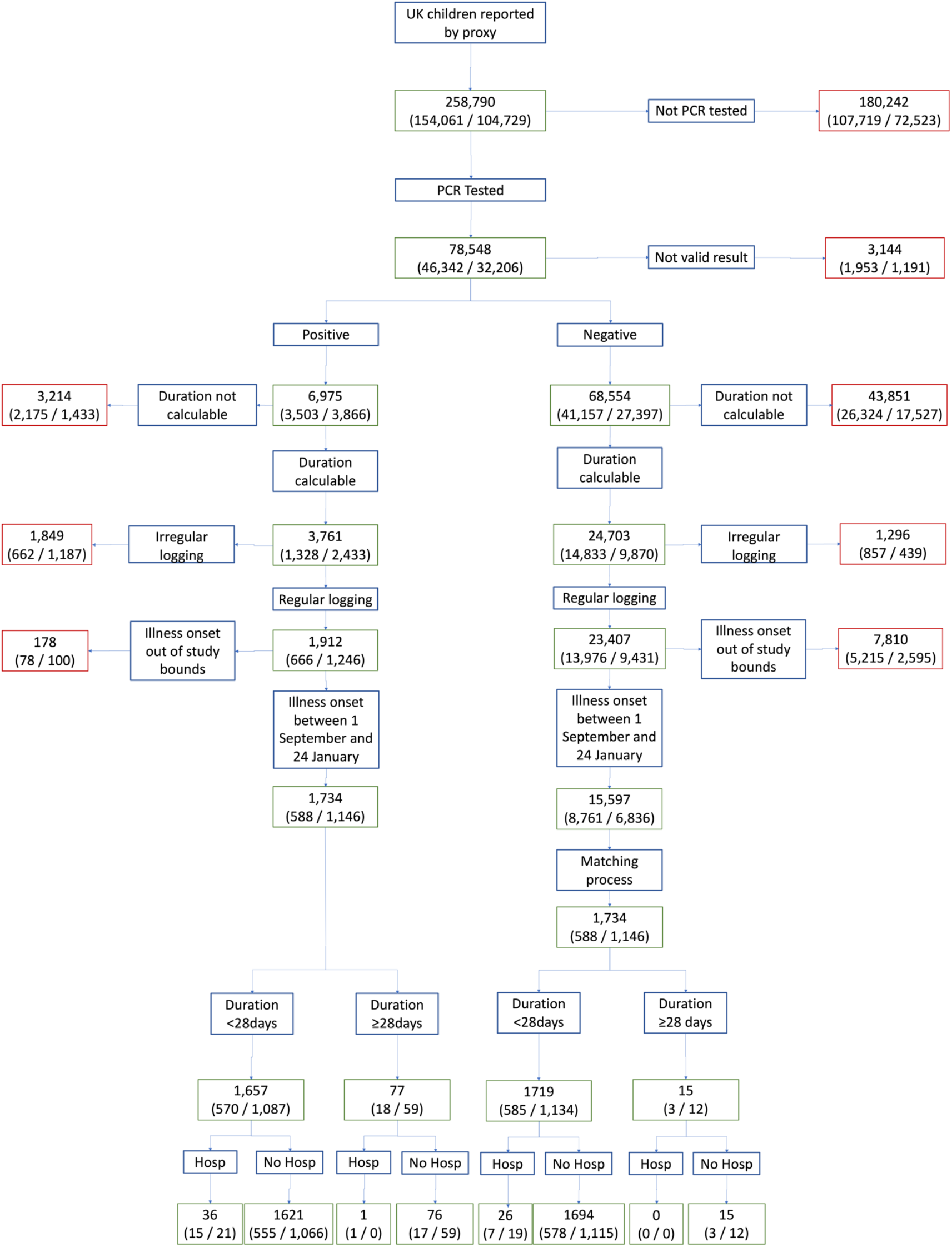
Flowchart of Inclusion and Exclusion Criteria for the Study. Legend: overall number for the entire cohort of children is given first; numbers within brackets separated by oblique refer to younger children and older children in that order. ‘Not valid result’ – PCR test result proxy-reported as “failed test” or “still waiting”. ‘Duration calculable’ – illness onset within defined timeframe of testing for SARS-CoV-2, and with defined endpoint (for details, please see Methods). ‘Irregular logging’ – proxy-reporting with intervals of >7 days between proxy-reports during illness duration). ‘Illness onset outside of study bounds’ - symptom onset before 1 September 2020 or after 24 January 2021. ‘Hosp’ – presenting to hospital (either admitted to hospital or seen in Emergency Department).

Among 16- and 17-year-old individuals, 29,047 self-logged data (447 reporting testing positive) compared to 32,271 proxy-reported (1,197 reported as testing positive). Illness duration could only be calculated in ten self-logged versus 381 proxy-logged 16- and 17-year-olds; and concurrent self-reporting and proxy-reporting could not be excluded. Thus, only proxy-reported data are presented here.

### Illness in children who tested positive for SARS-CoV-2

The median illness duration in children with COVID-19 was six [IQR 3;11] days (Table 1). Illness duration was shorter in younger compared with older children (five [IQR 2;9] vs. seven [IQR 3;12] days, Mann-Whitney U test p<1.e-5); and age correlated with illness duration (*r*_*s*_ 0.19, p<1.e-4).

**Table 1.** Characteristics of school-aged children who tested positive for SARS-CoV-2, and the control cohort of children (matched 1:1 for age, gender, and week of testing) who tested negative for SARS-CoV-2. The cohort of children with positive SARS-CoV-2 testing is presented here both as younger and older groups; and for usual (i.e., short) vs. extended illness duration.

|  | Children with positive SARS-CoV-2 test |  |  |  |  | Children with negative SARS-CoV-2 test (matched cohort) |
| --- | --- | --- | --- | --- | --- | --- |
|  | Younger group | Older group | Symptom duration <10 days | Symptom duration ≥ 28 days | Full cohort |  |
| **Number** | 588 | 1,146 | 1,183 | 77 | 1,734 | 1,734 |
| **Males (%)** | 287 (48.9) | 577 (50.3) | 618 (52.2) | 35 (45.5) | 864 (49.8) | 865 (49.9) |
| **Age, years (median, [IQR])** | 9 [7;10] | 15 [13;16] | 13 [10;15] | 14 [12;16] | 13 [10;15] | 13 [10;15] |
| **BMI (kg/m<sup>2</sup>) (median, [IQR])** | 17.0 [15.1;19.7] | 20.1 [17.8;22.3] | 19.0 [16.5;21.8] | 18.6 [16.3;21.8] | 19.2 [16.6;21.8] | 19.0 [16.5;21.5] |
| **Asthma (%)** | 69 (11.7) | 147 (12.8) | 134 (11.7) | 10 (13.0) | 216 (12.8) | 229 (13.2) |
| **Visit to hospital (%)** | 16 (2.7) | 21 (1.8) | 20 (1.7) | 1 (1.3) | 37 (2.1) | 26 (1.5) |
| **Illness duration**<br>**(median,**<br>**[IQR])** | 5 [2;9] | 7 [3;12] | 4 [2;6] | 46 [32;58] | 6 [3;11] | 3 [2;7] |
| **Number of**<br>**symptoms in**<br>**the first week**<br>**(median,**<br>**[IQR])** | 3 [2;5] | 4 [2;6] | 3 [2;5] | 6 [4;8] | 3 [2;6] | 2 [1;4] |

BMI: Body Mass Index. IQR: Inter Quartile Range.

Individual symptom prevalence and duration are shown in Table 2, Figure 2, and Figure 3. Overall, the most reported symptoms were headache (reported in 62.2% [55.1% younger, 65.9% older] children) and fatigue (55.0% [43.9% younger, 60.7% older] children). Subsequent symptoms ranked by frequency were in younger children: fever (43.7%), sore throat (36.2%), abdominal pain (27.7%), and persistent cough (24.7%); in older children: sore throat (51.0%), anosmia (48.3%), fever (34.6%), and persistent cough (26.0%). During first week of illness, the median symptom burden was three [IQR 2;6] overall (three [IQR 2;5] in younger, four [IQR 2;6] in older children).

**Table 2.** Symptoms over the duration of illness in children testing positive for SARS-CoV-2.

|  | **Sample tested positive for SARS-CoV-2** |  |  |
| --- | --- | --- | --- |
|  | **Younger children (aged 5-11 years, n=588)** | **Older children (aged 12-17 years, n=1,146)** | **Overall cohort (n=1,734)** |
| Headache | 324 | 755 | 1,079 (62.2%) |
| Fatigue | 258 | 696 | 954 (55.0%) |
| Sore Throat | 213 | 585 | 798 (46.0%) |
| Anosmia | 132 | 554 | 686 (39.6%) |
| Fever | 257 | 396 | 653 (37.7%) |
| Abdominal Pain | 163 | 194 | 357 (20.6%) |
| Dizziness | 84 | 300 | 384 (22.1%) |
| Persistent Cough | 145 | 298 | 443 (25.5%) |
| Loss of Appetite | 120 | 254 | 374 (21.6%) |
| Eye Soreness | 89 | 248 | 337 (19.4%) |
| Myalgias | 54 | 231 | 285 (16.4%) |
| Nausea | 95 | 193 | 288 (16.6%) |
| Hoarse voice | 63 | 166 | 229 (13.2%) |
| Chest Pain | 37 | 143 | 180 (10.4%) |
| Dyspnoea | 24 | 143 | 167 (9.6%) |
| Diarrhoea | 48 | 79 | 127 (7.3%) |
| Confusion | 15 | 81 | 96 (5.5%) |
| Red Welts | 16 | 36 | 52 (3.0%) |
| Blisters | 4 | 22 | 26 (1.5%) |

**Figure 2.**
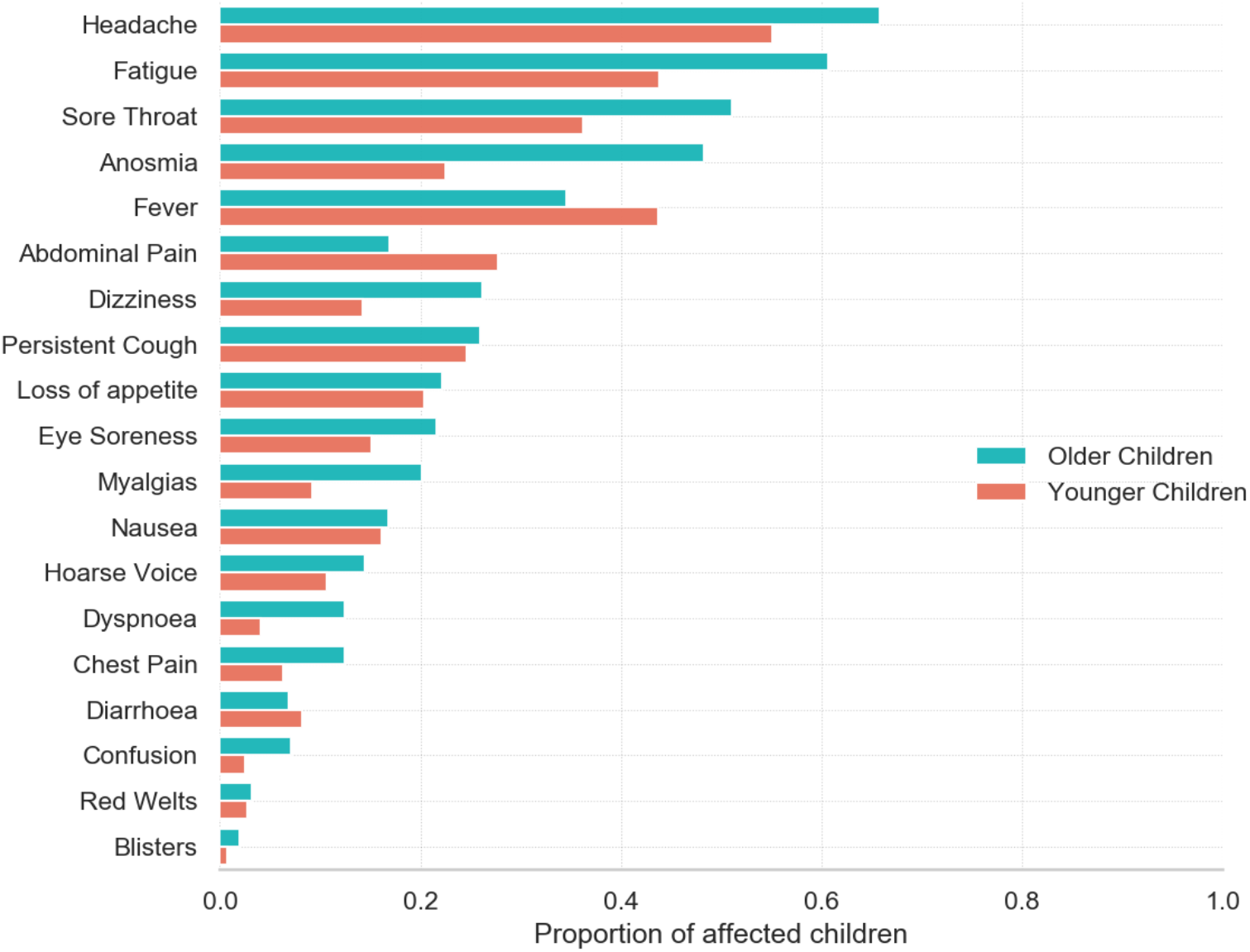
Prevalence of symptoms reported over the course of illness in younger (5-11 years, n=588) and older (12-17 years, n=1,146) children testing positive for SARS-CoV-2.

**Figure 3.**
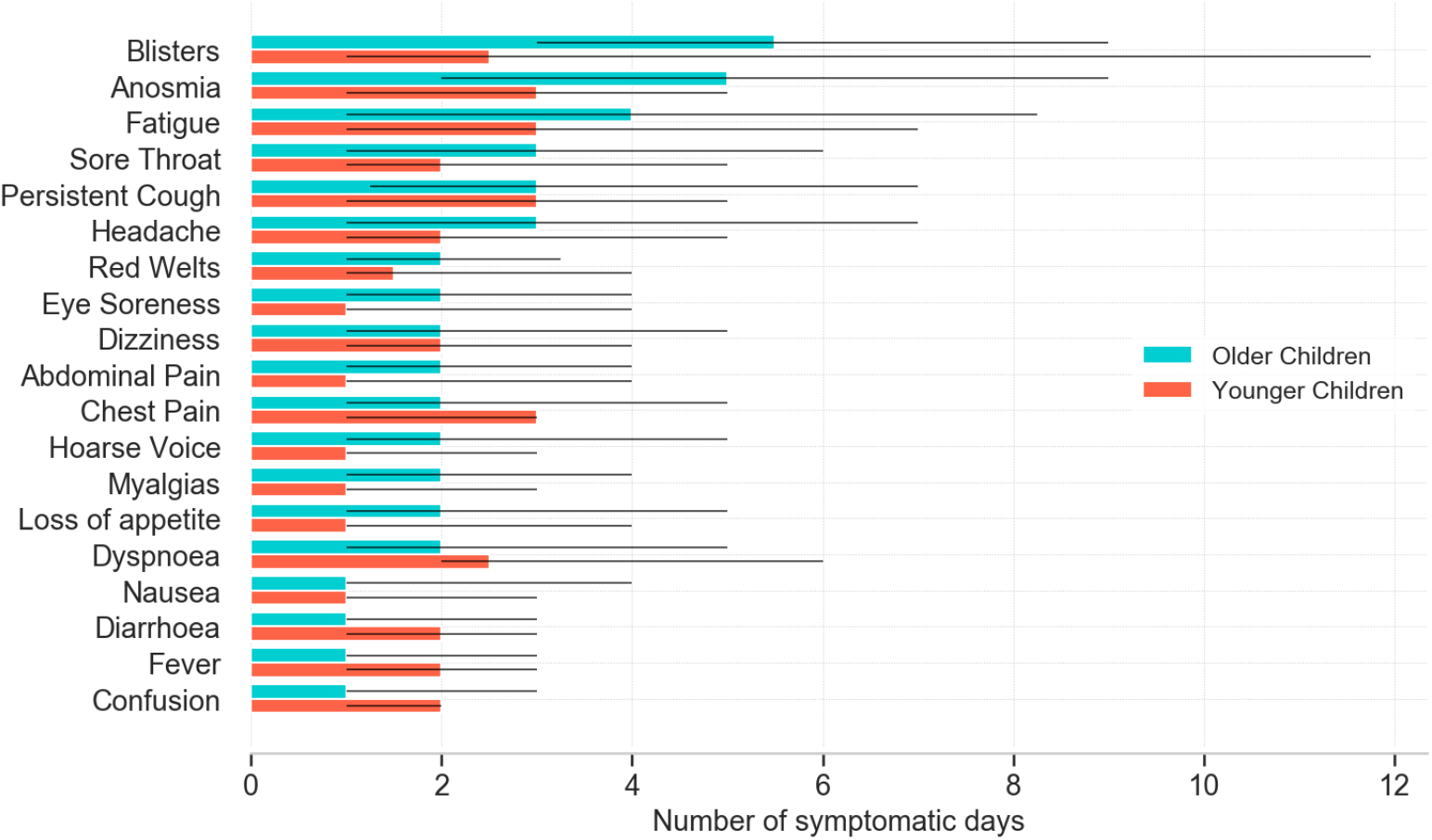
Median duration of each symptom [IQR] in younger (5-11 years) and older (12-17 years) children.

Sixteen younger and 21 older children testing positive for SARS-CoV-2 subsequently attended hospital. Symptom profiles of these children compared with children managed in the community are shown in Supplementary Figure 2. No formal statistical comparisons were undertaken between hospital and community cases, given the low numbers of hospital attendees.

### Long illness duration in children who tested positive for SARS-CoV-2

Overall, 77 (18 younger, 59 older) of 1,734 children (4.4% [95% CI 3.5-5.5]) had illness duration ≥28 days, meeting LC28 definition. The median symptom burden in these children was six symptoms [IQR 4;8] reported at least once during first week of illness, and eight symptoms [IQR 6;9] reported at least once over the entire illness duration. However, by day 28, median symptom burden was low, at two [IQR 1;4] symptoms (younger children: three [IQR 1;4]; older children, one [IQR 1;3]). The commonest symptoms experienced by these children over their entire illness were fatigue (experienced by 84.4%), headache (77.9%), anosmia (77.9%) and sore throat (74.0%). Figure 4 is a heat map of symptom profile and progression over the first 28 days in children with LC28.

**Figure 4.**
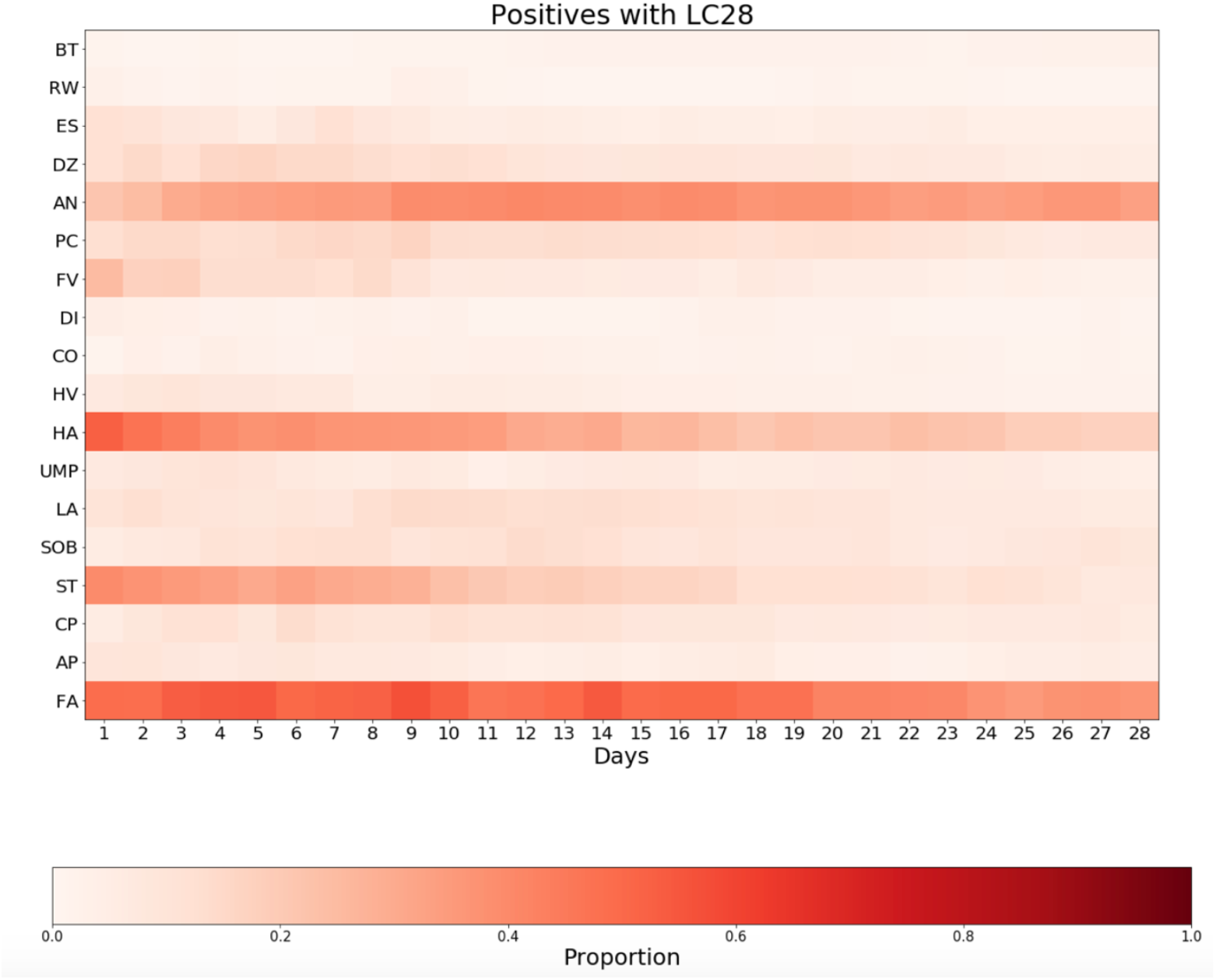
Heat maps showing symptom duration in school-aged children (aged 5-17 years) in whom at least one symptom persisted for ≥28 days. **Children with a positive SARS-CoV-2 test (n = 77 children)** Legend: X-axis, duration in days. Y axis, symptoms. Legend: BT, blisters; RW, red welts; ES, eye soreness; DZ, dizziness and light-headedness; AN, anosmia; PC, persistent cough; FV, fever; DI, diarrhoea; CO, confusion; HV, hoarse voice; HA, headache; UMP, myalgias [unusual muscle pains]; LA, loss of appetite; SOB, dyspnoea [shortness of breath]; ST, sore throat; CP, chest pain; AP, abdominal pain; FA, fatigue. Colour bar provides percentage comparison.

Twenty-five of 1,379 children (1.8% [95% CI 1.2-2.7]) had symptoms for ≥56 days, meeting LC56 definition.^3^ The median symptom burden in these children was six [IQR 4;8] symptoms reported at least once during the first week, and eight [IQR 6;10] symptoms reported at least once over the entire illness duration. The commonest symptoms experienced by these children over their entire illness were anosmia (84.0%), headache (80.0%), sore throat (80.0%) and fatigue (76.0%).

Older children were more likely to manifest symptoms ≥28 days compared with younger children (59 (5.1%) of 1146 older children vs. 18 (3.1%) of 588 younger children, Chi-squared two-tail test p=0.046). This was not significantly different in the smaller number of children with illness duration ≥56 days (19 (2.0%) of 934 older children vs. six (1.3%) of 445 younger children, Fisher’s exact test p=0.52).

### Symptom Reporting from Additional Questions (added 4 November 2020) and Free Text Analysis

Considering data from questions added to the app on 4 November 2020, the most reported symptom was rhinorrhoea (45.8% younger, 53.5% older children) then sneezing (36.3% younger, 36.6% older children) (Supplementary Figure 1), with similar prevalence in children with LC28 (51.4% rhinorrhoea, 48.9% sneezing) considered over their entire illness.

### Potential neurological symptoms

As mentioned, headache and fatigue were the commonest symptoms in children testing positive for SARS-CoV-2, overall and in each age group (Figure 2). Dizziness (undifferentiated between light-headedness and vertigo) was reported in 84 (14.3%) younger and 300 (26.2%) older children (median duration two [IQR 1;4] and two [IQR 1;5] days respectively). Symptoms of confusion (encompassing confusion, disorientation, and drowsiness) were reported in 15 (2.6%) younger and 81 (7.1%) older children (median duration two [IQR 1;2] and one [IQR 1;3] day(s) respectively). No formal statistical comparisons of these symptoms in children with shorter or longer illness duration were made, given low prevalence of these symptoms in children with LC28 (Figure 4).

Considering questions added on 4 November 2020 (Supplementary Table 2), with smaller sample size due to shorter timeframe, ‘brain fog’ was reported for 44 (8.7%) of 507 younger and 188 (20.2%) of 932 older children (median duration one day [IQR 1;4] and two days [IQR 1;5] respectively). Low mood was reported for 40 (8.0%) younger and 145 (15.6%) older children (median duration two [IQR 1;3.5] and two [IQR 1;4] days, respectively) (Supplementary Figure 1).

Free-text searching for specific neurologic symptoms disclosed very few reports of weakness (two children) or tics (one child). No severe neurological manifestations (paralysis, ataxia, epileptic seizures, fits, convulsions, paroxysms) were reported (Supplementary Table 3). Irritability (three children), emotional difficulties (two children), and behavioural difficulties (one child) were reported rarely; impaired attention and concentration were not reported.

### Illness in symptomatic children who tested negative for SARS-CoV-2

15,597 symptomatic children (8,761 younger, 6,836 older children) tested for SARS-CoV-2 but with a negative result, were proxy-reported at least once weekly within the requisite time frame and with calculable illness duration. Demographic details of the randomly selected matched control cohort of 1,734 children are shown in Table 1. Median illness duration in these children was three [IQR 2;7] days (younger children three [IQR 2;7] days; older children four [IQR 2;7] days), significantly shorter than for children testing positive for SARS-CoV-2 (Wilcoxon signed rank test p<1.e-5). The individual symptom duration is shown in Supplementary Figure 3 and prevalence in Supplementary Table 4. The most frequently reported symptoms over their entire illness were in younger children: sore throat (46.6%), headache (38.8%), fever (30.4%), fatigue (26.9%), and abdominal pain (24.7%); and in older children: sore throat (60.6%), headache (48.8%), fatigue (37.2%), fever (20.4%) and persistent cough (20.6%).

### Long illness duration in children who tested negative for SARS-CoV-2

Few children with a negative SARS-CoV-2 test had illness duration ≥28 days (15 of 1,734 children; 0.9%; 95% CI [0.5-1.4]), which was lower than for children testing positive considered overall (Chi-squared 2-tail test, p<1.e-10) and by age group (younger children: three (0.5%) vs. 18 (3.1%); Fisher’s exact test: p=0.001; older children, 12 (1.1%) vs. 59 (5.1%); Fisher’s exact test: p=0.001 and p<1.e-8). Symptom profile of these children over their first 28 days of illness is shown in Supplementary Figure 4. However, older children testing negative for SARS-CoV-2 with illness duration ≥28 days had a greater symptom burden than was experienced by children with LC28, both at ≥28 days (Supplementary Figure 5) and over their entire illness duration (Supplementary Figure 6) (p=0.005 and p=0.025 respectively, Mann-Whitney-U tests).

### Proxy-reporting density

Overall, proxy-reporting was assiduous for all children. Proxy-reporting density (number of logging episodes/illness duration in days) was significantly higher in children with a negative test for SARS-CoV-2, compared to children with a positive test (younger children: 1 [IQR 0.68;1] vs. 0.89 [IQR 0.6;1]; older children 1 [IQR 0.67;1] vs. 0.8 [IQR 0.57;1]), noting the shorter illness duration in children with a negative test.

### Proxy-reporting perseverance

Proxy-reporting until healthy was higher in children testing negative compared with children testing positive (1,674 of 1,734 [96.5%] vs. 1,551 of 1,734, [89.4%], Chi-squared 2-tail test p<1.e-15). Amongst all 1,734 children testing positive, logging ceased in 183 children prior to a healthy report (22 children with illness duration ≥28 days, 161 <28 days). In children with illness duration ≥28 days, proxy-reporting continued until a healthy report in 71.4% [55/77]. However, the remaining 22 children had already had symptoms for ≥28 days and thus fulfil the definition of LC28. In children with illness duration <28 days, a healthy report was received for 90.3% [1496/1657]. For the remaining 161 children, median symptom burden at last report was two [IQR 1;3]; and proxy-reporting usually ceased early in illness (logging cessation rates: 11.3% of children with illness duration <10 days; 5.5% ≥10 days). Thus, for children for whom proxy-reporting ceased prior to a healthy report, we have assumed that cessation corresponded to illness resolution (i.e., that proxy-reporting stopped because the child had recovered) and illness duration calculated accordingly.

We considered the impact of this assumption. Excluding all children testing positive but for whom a healthy report was not logged, median illness duration in the remaining 1,551 was unchanged (6 days [IQR 3;11]) and LC28 prevalence was 3.5% [55/1551], within the confidence intervals [3.5-5.5] for LC28 using data from the entire cohort. Excluding only the 161 children with [assumed] short symptoms but including all children with LC28 (regardless of receipt of healthy report), LC28 prevalence was 4.9% (77/1,573 children), again within the confidence intervals.

In children with a negative SARS-CoV-2 test, logging ceased in 60 children prior to a healthy report (four of 15 children with long illness duration, 56 of 1,719 children with [assumed] short illness duration).

As there was no data linkage between proxy-reported individual(s) and contributor, we cannot comment whether concurrent illness of family members affected proxy-reporting density or perseverance.

## Discussion

In this large study of UK school-aged children, we have shown that symptomatic SARS-CoV-2 infection in children is usually of short duration, with median illness duration of six days (compared to eleven days in adults^3^) and with low symptom burden. Prolonged illness duration can occur but is infrequent (4.4% ≥28 days; 1.8% ≥56 days), lower than observed in adults using the same disease definitions (in adults: prevalence of LC28, 13.3% and LC56, 4.5%).^3^ Age correlated with illness duration (overall, and in children with illness duration ≥28 days), consistent with our previous findings in adults.^3^

Similar to adults,^3^ the commonest symptoms in children with COVID-19 were headache (62.2%) and fatigue (55.0%) (Table 2, Figure 2). A meta-analysis of studies including both community-based and hospitalized children with COVID-19 identified fever (47%) and cough (42%) as the commonest symptoms; however, headache and fatigue were only assessed in half the contributing cohorts.^4^ In our cohort, fever was reported for 37.7% and persistent cough in 25.5%. Anosmia (including anosmia and dysosmia) was common (39.6% overall), and higher than observed in a small study of adolescents aged 10-19 years with ‘mild to moderate’ COVID-19 (here, anosmia: 24.1%),^17^ noting that anosmia was one of the core symptoms determining access to testing in the UK during our study period.

In children with illness duration ≥28 days, by day 28 symptom burden was low (median, two). Fatigue was proxy-reported at some stage in almost all (84.4%) of these children (Figure 4, Supplementary Figure 6). Fatigue has been reported as the commonest symptom of long COVID in many adult studies, although prevalence varies. In our own adult study, fatigue was almost universal in LC28 (97.7% at some stage during their illness);^3^ other adult studies have reported persistence of fatigue as 53.1% at 60 days;^18^ and 52.3% at 10 weeks.^19^

Few large-scale epidemiological studies have been undertaken to provide normative data of prevalence and persistence of headache and fatigue in the general paediatric population. Regarding headache, a systematic review of 38 unselected population-based studies in individuals aged <20 years reported ∼60% were “prone to headache”.^20^ A random sample of 2,165 school children aged 5-15 years reported 1,166 (66%) had headaches over the previous year.^21^ Regarding fatigue, a study of 2,936 children found 129 (4.4%) had “more than a few days of disabling fatigue”.^22^ Prevalence of chronic fatigue syndrome (“disabling fatigue lasting >3 months…where no other cause for the fatigue could be established”) was 1% of children aged 11-16 years in a study from three English secondary schools;^23^ and prevalence of “chronic disabling fatigue” (“fatigue lasting >6 months associated with absence from full-time school or that had prevented…[participation] in activities”) in the Avon Longitudinal Study was 1.5% and 2.2% in children aged 13 and 16 years respectively.^24^ Considering fatigue after viral infection, median illness duration after Epstein–Barr virus in symptomatic university students was ten days (mean 17, range 3-66 days), with fatigue persisting for a median of 15.5 days.^25^ These studies indicate the wide range of prevalence of these symptoms in paediatric populations generally.

Lack of contemporaneous data comparing illness duration and symptom profiles after infection with different viruses also complicates contextualisation of COVID-19 in children. A preprint paper comparing data from 55,270 children/adolescents with COVID-19 (3,693 of whom were hospitalized) with a non-contemporaneous cohort of 1,952,693 children with influenza during 2017-19 (hospitalised numbers unclear) suggested that dyspnoea, anosmia, and gastrointestinal tract symptoms were more common in children with COVID-19 than in children with influenza.^26^ However, symptoms and overall illness duration were only reported as present or absent at 30 days in both groups, preventing more granular comparisons.

A strength of our study is our comparison of contemporaneous illness profiles of symptomatic children testing positive vs. negative for SARS-CoV-2, matched for age, gender, and week of testing. Children testing positive had longer median illness duration (six days vs. three days in children testing negative); and were more likely to have illness duration ≥28 days (4.4% vs. 0.9%). However, some children testing negative also had illness duration ≥28 days, and these children had a higher symptom count both over their entire illness duration and at day 28, acknowledging here that our sample size is small. We considered whether some of these children might have false negative results. However, there is no evidence that sensitivity and specificity of SARS-CoV-2 testing differ in children compared with adults, noting sensitivity for PCR SARS-CoV-2 testing of ∼95%.^27^ The symptom profiles of these children (Supplementary Data) suggest some differences compared with children testing positive, though this could not be assessed statistically. Relevantly, prevalence of non-SARS-CoV-2 respiratory viruses (influenza A, influenza B, parainfluenza, adenovirus, rhinovirus, and respiratory syncytial virus) were unusually low over the 2020-21 UK winter,^14,28,29^ with the exception of the rhinovirus peak commonly observed in September with return-to-school.^14^ With relaxation of personal protection and social distancing, these illnesses are likely to return towards more usual (i.e., higher) incidences in future UK winters. Our data highlight that other illnesses may also have a protracted and burdensome course in children: this needs to be considered in post-pandemic service delivery planning.

Short and long-term effects of COVID-19 on school performance and learning have been a matter of concern.^30^ In our cohort, neither attentional problems nor memory complaints nor anxiety were reported. Isolated cases of low mood and irritability were consistent with previously reported figures in the school-aged healthy population.^31^ Our data do not support anecdotal reports of weakness and seizures as common features in children with COVID-19, whether of short or long illness duration; and no severe neurological symptoms were reported. However, symptom persistence from any illness can cause low mood, with adverse long-term outcomes including school refusal and separation anxiety.^32^ Although self-reporting adults could participate in specific questions regarding mental health, these data could not be proxy-reported; this limits our ability to assess mental health comprehensively in children during the pandemic, whether testing positive or negative for SARS-CoV-2.

In considering our data relative to other sources, the UK Office for National Statistics (ONS) conducted a round of SARS-CoV-2 testing (irrespective of symptoms) from 2-10 December 2020 (i.e., prior to the UK peak), in 7,089 pupils drawn from 121 schools (41 primary, 80 secondary); positive tests were reported in 0.94% of primary school pupils and 0.99% of staff, and in secondary schools 1.22% of pupils and 1.64% of staff. However, the ONS warned that their conclusions might not have general validity, as there was deliberate oversampling of schools that had higher infection rates early after return-to-school.^33^ Moreover, these figures do not capture fluctuations as the pandemic progressed. In our dataset integrating data across the pandemic, 6,975 (2.7%) of 258,790 of proxy-reported children had a positive test (2.3% of younger, 3.7% of older children), noting that: a) testing was only available for symptomatic individuals, and rarely during the first wave; and b) proxy-reporting was voluntary and through a COVID-19 study app, with its (adult) user base over-representative of females, white background, and above-average socioeconomic status compared with the general UK population.^12^ Further, we cannot characterise regional variability, as geographic information was unavailable for many participants.

We considered the proportion of all UK school-aged children testing positive for SARS-CoV-2 proxy-reported in our study. Different UK countries report data for children within different time periods and different age groups (e.g., often including young adults aged 18-19 years and preschool-aged children) (Supplementary Table 5). Conservatively, our study represents ∼1-2% of UK school-aged children testing positive during the pandemic.

The most recent ONS data (released April 2021) estimated 9.8% of children aged 2-11 years and 13.0% of children aged 12-16 years experiencing ongoing symptoms five weeks after a positive SARS-CoV-2 testing, with 7.4% and 8.2% respectively still reporting symptoms at 12 weeks.^34^ These figures were a reduction compared with previous ONS estimates in January 2021 (e.g., from 12.9% to 9.8% in children aged 2-11 years).^34,35^ The ONS has reported a control group (defined as never symptomatic, never tested, never self-isolated, and never a contact of anyone testing positive), with ‘baseline’ symptom rates (not otherwise parsed) of 2% in children aged 2-11 years and 1.7% in children aged 12-16 years.^34,35^ There is limited published detail of ONS methodology.

Part of the disparity between our LC28 prevalence in children and the ONS figures may be that the ONS required two consecutive asymptomatic visits to define illness end: thus, children with asymptomatic periods >1 week between symptoms would be captured by ONS but not by our study. Notably, the ONS’ sensitivity analysis of the impact of defining illness end to a single asymptomatic visit markedly lowered their prevalence estimates, especially for illness >12 weeks (from 13.7% to 0.9%^34^), more consistent with our results. Additionally, ONS estimates use current and recalled data collected in the first week of each month, whereas our app-based data were collected in real time with high proxy-reporting density and persistence. Our data concords with a small Australian study reporting 171 young children testing positive for SARS-CoV-2 (median age three years [IQR 1;8]), 151 of whom were followed for 3-6 months: twelve (8%) children (median age, two years) had symptoms 3-8 weeks after initial presentation (most commonly, cough and/or fatigue); all returned to baseline health by study end.^36^

Our study is part of one of the largest citizen-scientist initiatives ever in the UK. We leveraged our previously published methodologies assessing illness duration and symptom profiling, including long COVID assessment.^3^ Our data census points allowed us to capture all children with illness duration ≥8 weeks who presented before the UK peak positive specimen date, and by ensuring symptom presentation concorded with test timing we could attribute symptoms to illness defined by contemporaneous test results. We avoided bias from limited test availability early in the pandemic by restricting analyses to start from 1 September 2020. We acknowledge there were still some blocks to testing access^37^ – specifically, individuals were required to have fever, cough, or anosmia which symptom list was largely informed by adult symptomatology and might not capture some common paediatric manifestations of COVID-19 (e.g., abdominal pain, reported in 4% of paediatric cases^4^ and in 27.8% of our younger children). Similarly, direct questions asked through the app were largely informed by research in adults. The free-text entries did not suggest common symptoms unique to paediatric populations; however, we did not undertake a formal qualitative analysis of free-text data given (a) its *ad hoc* collection and (b) the potential bias arising from introduction of additional direct questions from 4 November 2020 (i.e., once a symptom was directly asked, it was unlikely to be reported as free-text). We also acknowledge that symptoms were proxy-reported rather than directly ascertained; however, this is common in clinical assessment of children, particularly younger children.

## Conclusions

Our national study provides the first systematic description of COVID-19 in symptomatic school-aged children tested for SARS-CoV-2. Our data show that long illness duration after COVID-19 is uncommon; however, a small proportion of children do have prolonged illness duration and persistent symptoms, validating these children’s experiences. Our LC56 data provide reassurance regarding longer outcome for these children. The symptom burden in children testing negative for SARS-CoV-2 but with long illness duration highlights that allocation of appropriate resources will be necessary for any child with prolonged illness, whether from SARS-CoV-2 infection or not. Our study provides timely and critical data to inform discussions around the impact and implications of the pandemic on UK paediatric healthcare resource allocation.

## Data sharing

Data collected in the COVID Symptom Study smartphone application are being shared with other health researchers through the UK National Health Service-funded Health Data Research UK (HDRUK) and Secure Anonymised Information Linkage consortium, housed in the UK Secure Research Platform (Swansea, UK). Anonymised data are available to be shared with researchers according to their protocols in the public interest (https://web.www.healthdatagateway.org/dataset/fddcb382-3051-4394-8436-b92295f14259).

## Authors contributions

EM, CHS, LSC, ELD performed the analyses

BM, EK, LC, JD, MM performed data extraction and curation

EM, SSB, RCH, AH, MA, ELD wrote the manuscript

CH, SSe, KR, JCP developed the data collection system

TDS, SO, CJS conceived the CSS and obtained funds

MM, SO, CJS, ELD coordinated this research

All the authors critically reviewed the manuscript

## Ethics Committee Approval

Ethics approval for this study was granted by the KCL Ethics Committee (reference LRS-19/20-18210); all participants (here, proxy-reporting adults) provided consent.

