## Supplementary Material for "Illness duration and symptom profile in a large cohort of symptomatic UK school-aged children tested for SARS-CoV-2"

**Supplementary Table 1. List of symptom questions asked by the COVID Symptom Study app at 1 September 2020. Answers were yes/no, unless indicated otherwise.**

| Symptom | COVID Symptom Study app question |
|---|---|
| Fever | Do you have a fever? |
| Persistent Cough | Do you have a persistent cough (coughing a lot for more than an hour, or 3 or more coughing episodes in 24 hours)? |
| Fatigue | Are you experiencing unusual fatigue? (no; mild fatigue; severe fatigue/ I struggle to get out of bed) |
| Dyspnoea | Are you experiencing unusual shortness of breath? (no; yes mild symptoms/ slight shortness of breath during ordinary activity: yes significant symptoms - breathing is comfortable only at rest; yes, severe symptoms/ breathing is difficult even at rest). |
| Anosmia | Do you have a loss of smell/taste? |
| Hoarse Voice | Do you have an unusually hoarse voice? |
| Chest Pain | Are you feeling an unusual chest pain or tightness in your chest? |
| Abdominal Pain | Do you have an unusual abdominal pain? |
| Diarrhoea | Are you experiencing diarrhoea? |
| Headache | Do you have a headache? How often are you experiencing headaches? (number) |
| Confusion | Do you have any of the following symptoms: confusion, disorientation or drowsiness? |
| Eye Soreness | Do your eyes have any unusual eye-soreness or discomfort (e.g., light sensitivity, excessive tears, or pink/red eye)? |
| Loss of Appetite | Have you been skipping meals? |
| Nausea | Have you felt nauseous or experienced vomiting? |

| | |
|---|---|
| **Dizziness** | Are you experiencing dizziness or light-headedness? |
| **Sore Throat** | Do you have a sore throat? |
| **Myalgias** | Do you have unusual strong muscle pains? |
| **Red Welts** | Have you had raised, red, itchy welts on the skin or sudden swelling of the face or lips? |
| **Blisters** | Have you had any red/purple sores or blisters on your feet, including your toes? |

**Supplementary Table 2. List of symptom questions asked by the COVID Symptom Study app after changes implemented on 4 November 2020.** Questions were: Do you have (symptom)? Answers were yes/no, unless indicated otherwise. New symptoms added on 4 November 2020 are indicated in bold.

| **Symptom** | **COVID Symptom Study app question** |
|---|---|
| Fever | Fever (at least 37.8C or 100F) |
| Persistent Cough | Persistent cough (coughing a lot for more than an hour or 3 or more coughing episodes in 24 hours) |
| Fatigue | Unusual fatigue… (no; mild fatigue; severe fatigue/ I struggle to get out of bed) |
| Dyspnoea | Shortness of breath or trouble breathing (no; yes mild symptoms/ slight shortness of breath during ordinary activity: yes significant symptoms/ breathing is comfortable only at rest; yes, severe symptoms/ breathing is difficult even at rest). |
| Anosmia**/Ageusia** | Loss of smell / taste |
| Hoarse Voice | Unusually hoarse voice |
| Chest Pain | Unusual chest pain or tightness in your chest |
| Abdominal Pain | Unusual abdominal pain or stomach-ache |
| Diarrhoea | Diarrhoea |
| **Stools** | How many loose stools in the last 24 hours? |
| **Headache Frequency** | How often are you experiencing headaches? |
| Confusion | Confusion, disorientation or drowsiness |
| Eye Soreness | Do your eyes have any unusual eye-soreness or discomfort (e.g. light sensitivity, excessive tears, or pink/red eye)? |
| Loss of appetite | Skipping meals |

| | |
|---|---|
| Headache | Headache |
| Nausea | Nausea or vomiting |
| Dizziness | Dizziness or light-headedness |
| Sore Throat | Sore or painful throat |
| Myalgias | Unusual strong muscle pains or aches |
| Red Welts | Raised, red, itchy welts on the skin or sudden swelling of the face or lips |
| Blisters | Red/purple sores or blisters on your feet, including your toes |
| **Allergy Exacerbation** | Increase in your usual allergy symptoms |
| **Rashes** | Rash on your arms or torso |
| **Sensitive Skin** | Strange, unpleasant sensations in your skin like pins & needles or burning |
| **Hair Loss** | Unusual hair loss |
| **Low Mood** | Feeling down, depressed or hopeless |
| **Brain Fog** | Loss of concentration or memory (brain fog) |
| **Dysosmia/Dysgeusia** | Altered smell / taste (things smell or taste different to usual) |
| **Rhinorrhoea** | Runny nose |
| **Sneezing** | Sneezing more than usual |
| **Ear Pain** | Earache |
| **Tinnitus** | Ringing in your ears |
| **Lymphadenopathy** | Swollen neck glands |

| | |
|---|---|
| **Palpitations** | Unusually fast or irregular heartbeat (palpitations) |
| **Arthralgias** | Unusual joint pains or aches |
| **Mouth Ulcers** | Mouth or tongue ulcers |
| **Tongue Changes** | Changes to tongue surface |

**Supplementary Table 3. Theme summary from the free-text scrutiny.**

Twelve themes were identified and ordered by text word frequency. For each theme, numbers of children (overall and within younger/older age groups) are charted, with keywords used in the automatic search (asterisk at the end of search keywords indicates free term termination (wildcard)) and the number of children for whom each simple or combined term was reported. Free-text was also searched for specific terms of interest, including neuromuscular symptoms (e.g., weakness, paralysis, tics, seizure), and symptoms potentially affecting attention, learning, and school performance (e.g., anxiety, irritability). Summation of the number of children with each symptom within a theme may not equal the number of children within the theme overall, due to multiple terms and negations in free-text.

| Theme number | Theme name | Number overall | Number of younger children | Number of older children | Search keywords | Numbers of individuals reporting a term |
|---|---|---|---|---|---|---|
| 1 | Respiratory tract symptoms | 131 | 41 | 90 | nose, sinus, chest, congest*, cold, sneez*, wheez*, throat, tonsillitis, asthma, phlegm, mucus, coughing blood | cold (46), blocked nose, blocked sinuses (17), stuffy nose (5), sinus pain, burning sinuses, sore nose, nose pain (18), congestion (12), burning nose (3), clearing throat (7), dry throat (1), tickle in throat, irritated throat, itchy throat (6), tonsillitis (5), headcold (1), sneezing (2), nose bleeding (2), mucus (8), phlegm (11), fleming (2), asthma (8), wheeze (1), quick breath (1), tight chest (2), burning sensation in chest (1), "pins and needles" in chest (1). |
| 2 | Cutaneous manifestations | 42 | 18 | 24 | skin, rash, eczema, impetigo, pale, itch*, spot, finger | eczema (7), rash (5), body rash (1), rash on torso (3), rash on hands (1), rash on face (7), rash on elbows (1), sensitive skin (4), itchy skin (9), itchy bumps on toe skin, toe lumps (2), dry |

| | | | | | | |
|---|---|---|---|---|---|---|
| | | | | | | skin (2), red spots (3), white spots on legs (1), finger chilblains (3), pale (4), chilblain on finger knuckle (1). |
| 3 | Oral cavity manifestations | 29 | 11 | 18 | mouth, tongue, lip, throat ulcer | mouth ulcers (6), sore or sensitive mouth (4), tongue rash (3), tied tongue and difficulty in speaking (1), white or purple spots on tongue (2), white tongue (1), dry mouth (1), dry lips (1), swollen lips (1), burning or sore lips (3), rash around the lips (1). |
| 4 | Joint symptoms and lymphadenopathy | 23 | 9 | 14 | gland, sore | swollen glands (3), sore legs or hip (2), sore back (1), sore joints (1), sore neck glands (1). |
| 5 | Ocular symptoms | 16 | 2 | 14 | eye, vision, teary | aching or sore eyes (1), itchy eyes (1), pain behind the eyes (1), eye heaviness or discomfort (3), blurred vision (2), double vision from one eye (2), seeing purple (2), twitchy eye (1), teary eyes (1). |
| 6 | Cardiovascular/autonomic symptoms | 14 | 4 | 10 | sweaty, faint, flush, palpitations, heartbeat, shak* | shaky (2), faint (2), palpitations (1), rapid intermittent heartbeat (1), flushed (1), sweaty (1). |
| 7 | Sleep disturbance | 13 | 3 | 10 | sleep, insomnia | difficulty or problems in sleeping (5), disturbed or broken sleep (5), unusual moaning in sleep (1), day sleepiness (1), unable to sleep (1). |
| 8 | Otological symptoms | 7 | 1 | 6 | ear | hearing loss (1), ringing or popping in ears (2), blocked ear (1), itchy ear (1), glue ear (1). |
| 9 | Mental, mood and affective health | 6 | 4 | 2 | concentration, attention, focus, irritab*, emotion, | irritability (2), low mood (1), “very grumpy” (1), emotional (1), odd behaviour (1). |

| | | | | | | |
|---|---|---|---|---|---|---|
| | | | | | grumpy, behav*, mood, anxiety | |
| 10 | Neurological | 6 | 1 | 5 | tic, twitch, weak*, paralysis, balance, ataxia, walk*, seizure, convulsions, paroxysm, sensory, fit | physical tics (1), weakness (2). |
| 11 | Genitourinary symptoms | 5 | 3 | 2 | urin*, bladder, kidney, penis, genital itching | frequent need to urinate (1), urinary tract infection (2), penis infection (1). |
| 12 | Gastrointestinal symptoms | 3 | 1 | 2 | constipation, stomach | stomach discomfort (2). |
| 13 | Miscellaneous | 17 | 6 | 11 | jaw, thirst, swelling, sugar, toe | swelling (2), low blood sugar with no diabetes (1), increased thirst (2), mottled toes (1), toe swelling (1), toe lumps (1). |

**Supplementary Table 4. Number of subjects reporting each symptom over the course of illness in younger (5-11 years, n=588), older (12-17 years, n=1,146) and overall (n=1,734) children testing negative for SARS-CoV-2.**

| | **Sample tested negative for SARS-CoV-2 test** | | |
|---|---|---|---|
| | **Younger children (aged 5-11 years, n=588)** | **Older children (aged 12-17 years, n=1,146)** | **Overall cohort (n=1,734)** |
| Headache | 228 | 559 | 787 (45.4%) |
| Fatigue | 158 | 426 | 584 (33.7%) |
| Sore Throat | 274 | 695 | 969 (55.9%) |
| Anosmia | 45 | 142 | 187 (10.8%) |
| Fever | 179 | 234 | 413 (23.8%) |
| Abdominal Pain | 145 | 176 | 321 (18.5%) |
| Dizziness | 43 | 193 | 236 (13.6%) |
| Persistent Cough | 137 | 236 | 373 (21.5%) |
| Loss of Appetite | 67 | 148 | 215 (12.4%) |
| Eye Soreness | 37 | 101 | 138 (8.0%) |
| Myalgias | 34 | 114 | 148 (8.5%) |
| Nausea | 90 | 219 | 309 (17.8%) |
| Hoarse voice | 68 | 185 | 253 (14.6%) |
| Chest Pain | 21 | 104 | 125 (7.2%) |
| Dyspnoea | 21 | 70 | 91 (5.2%) |
| Diarrhoea | 71 | 88 | 159 (9.2%) |
| Confusion | 8 | 40 | 48 (2.8%) |
| Red Welts | 19 | 20 | 39 (2.2%) |
| Blisters | 8 | 20 | 28 (1.6%) |

**Supplementary Table 5.** Official data of SARS-CoV-2 positive cases in children and young adults in England, Scotland, Wales and Northern Ireland.

| | **Number of positive tests** | **Age [years]** | **Period** | Number | Age [years] |
|---|---|---|---|---|---|
| **England**[1] | 390,866 | 5-19 | 1 September 2020 to 24 January 2021 | 69,641 | 5-9 |
| | | | | 321,225 | 10-19 |
| **Scotland**[2] | 30,466 | 0-19 | Start of COVID-19 pandemic to 30 March 2021 | 15,869 | 0-14 |
| | | | | 14,597 | 15-19 |
| **Wales**[3] | ~28,300 | children and young people | Start of COVID-19 pandemic to 30 March 2021 | - | - |
| **Northern Ireland**[4] | 13,268 | 0-19 | Start of COVID-19 pandemic to 30 March 2021 | - | - |

**Supplementary Figure 1. Proportions of children with a positive test for SARS-CoV-2 with symptoms from questions added to the app, from 4 November 2020.**

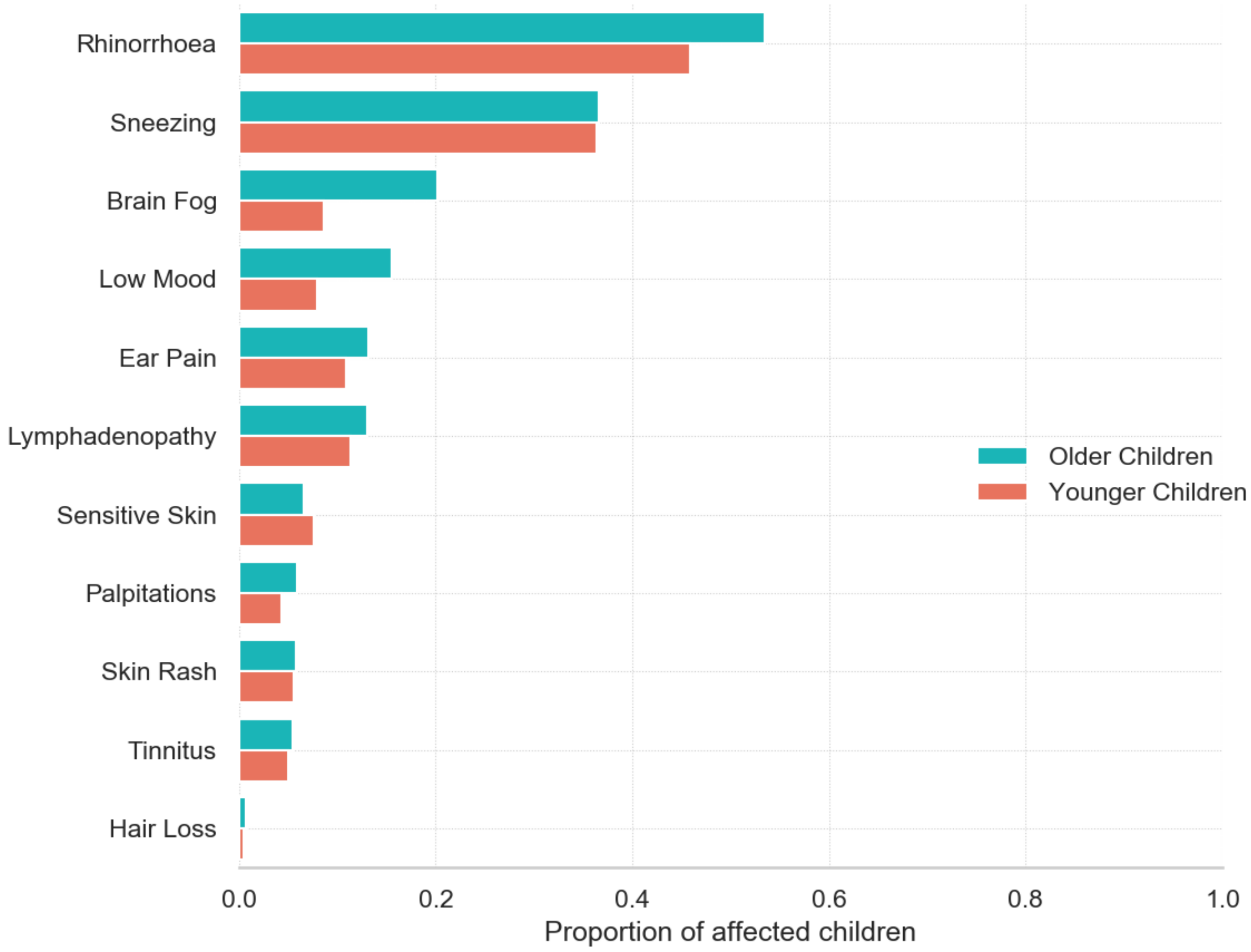

**Supplementary Figure 2. Individual symptom profile and prevalence in younger children (YC, left panel) and older children (OC, right panel) testing positive for SARS-CoV-2, comparing 37 (16 younger, 21 older) children presenting to hospital (darker bars) with children managed in the community (lighter bars).**

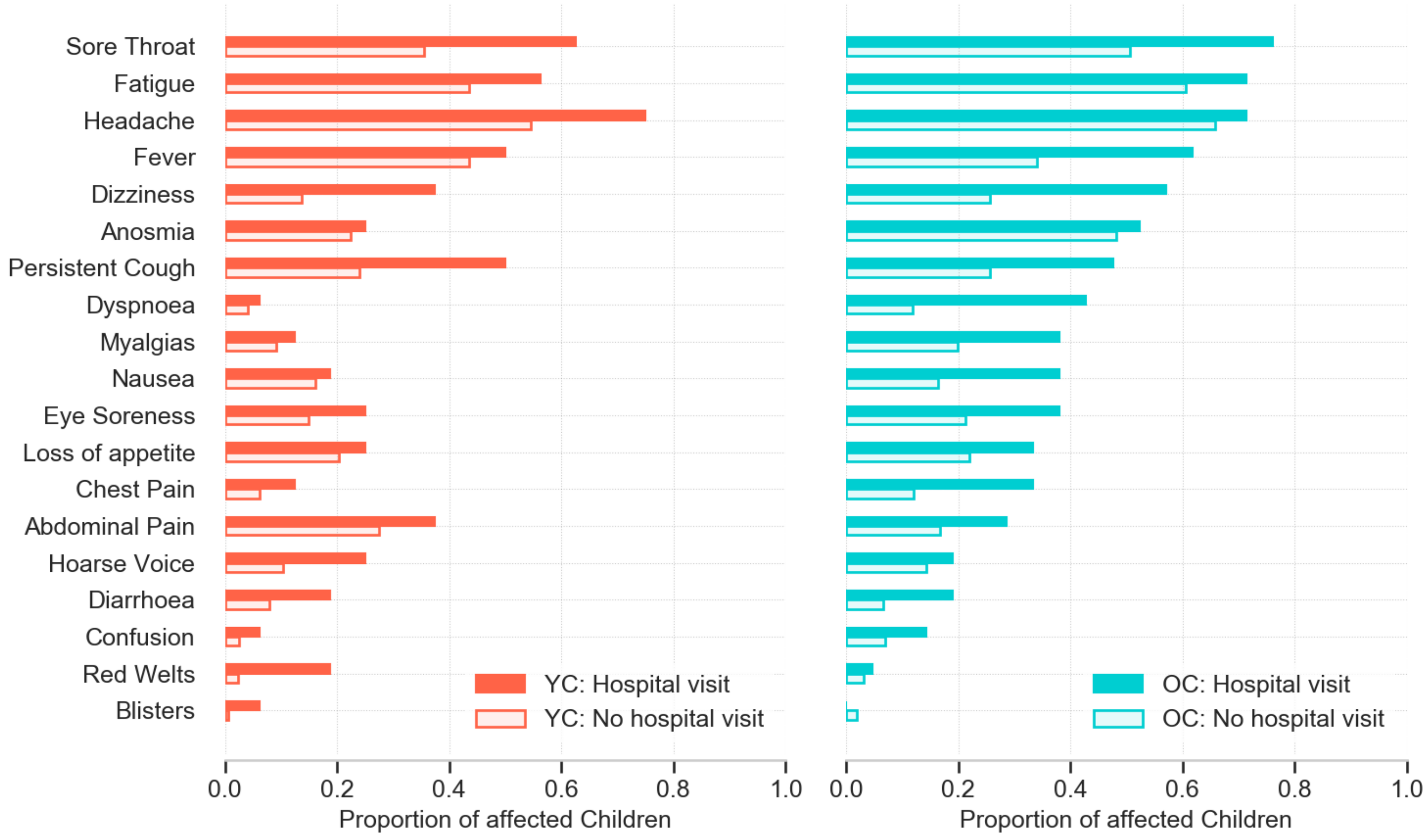

**Supplementary Figure 3. Symptom duration [IQR] in the control cohort (matched for age, gender, and week of testing) in younger (5-11 years) and older (12-17 years) children negative for SARS-CoV-2 (n=1734).**

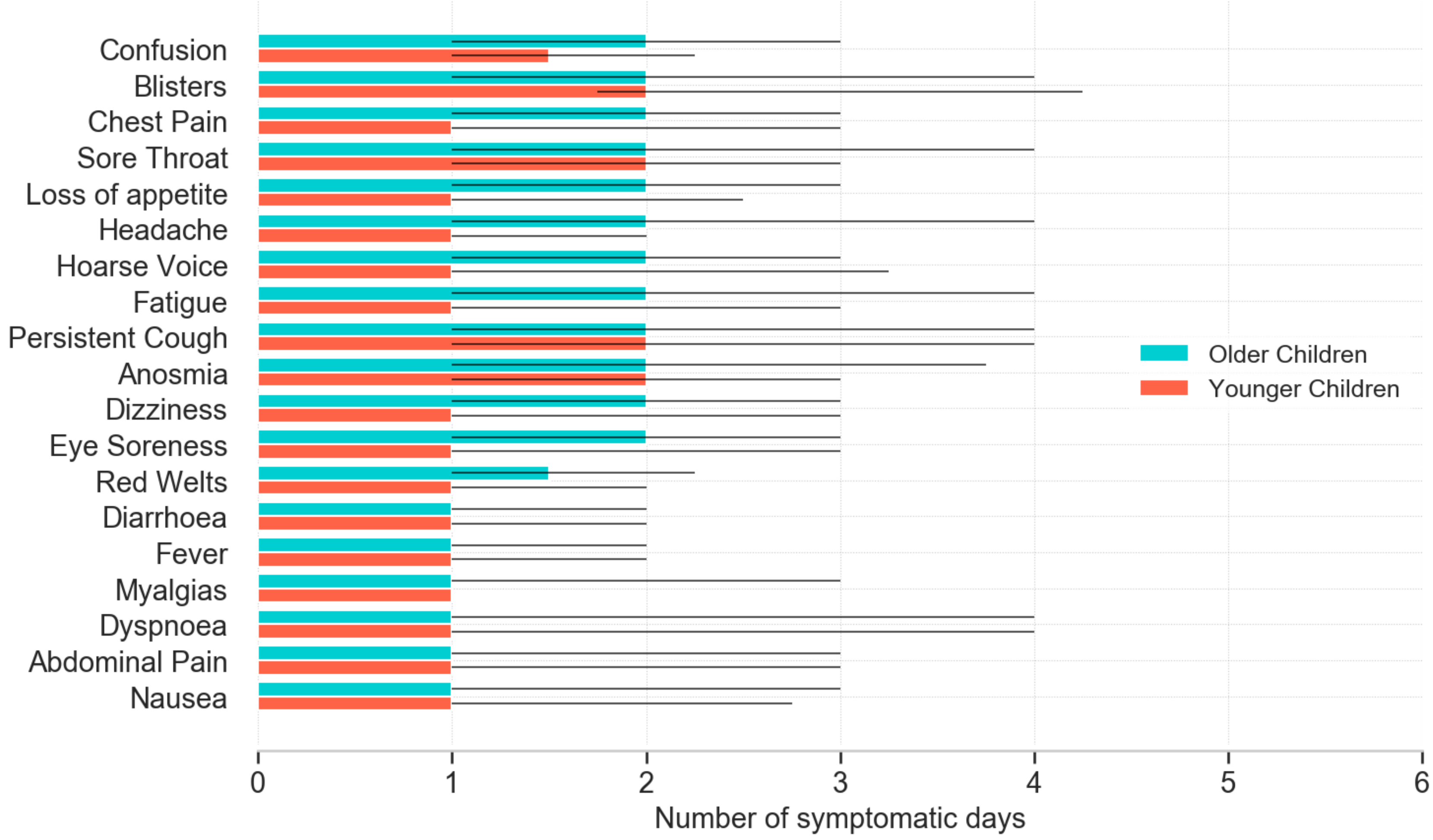

**Supplementary Figure 4. Heat map showing symptom duration in children (aged 5-17 years) with a negative SARS-CoV-2 test,** in whom at least one symptom persisted for **≥28 days (n =15 children).**

Legend:

X-axis, duration in days.

Y axis, symptoms.

Legend: BT, blisters; RW, red welts; ES, eye soreness; DZ, dizziness and light-headedness; AN, anosmia; PC, persistent cough; FV, fever; DI, diarrhoea; CO, confusion; HV, hoarse voice; HA, headache; UMP, myalgias [unusual muscle pains]; LA, loss of appetite; SOB, dyspnoea [shortness of breath]; ST, sore throat; CP, chest pain; AP, abdominal pain; FA, fatigue. Colour bar provides percentage comparison.

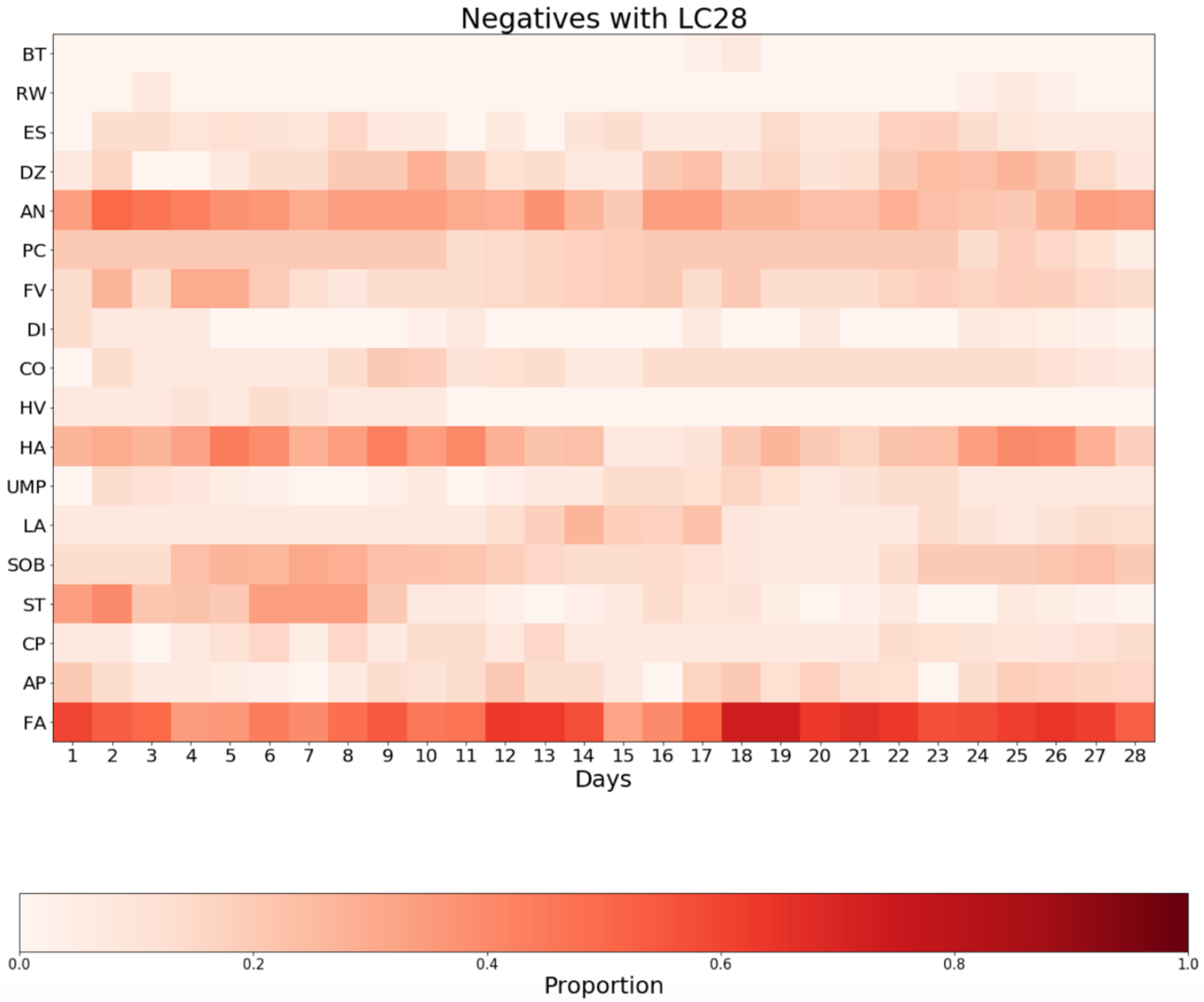
Negatives with LC28
BT
RW
ES
DZ
AN
PC
FV
DI
CO
HV
HA
UMP
LA
SOB
ST
CP
AP
FA
1 2 3 4 5 6 7 8 9 10 11 12 13 14 15 16 17 18 19 20 21 22 23 24 25 26 27 28
Days
0.0 0.2 0.4 0.6 0.8 1.0
Proportion

**Supplementary Figure 5. Symptom profile (at day 28 or beyond) in younger children (YC, left panel) and older children (OC, right panel) with illness duration ≥28 days. Each panel compares children positive (darker bars) and negative (lighter bars) for SARS-CoV-2.**

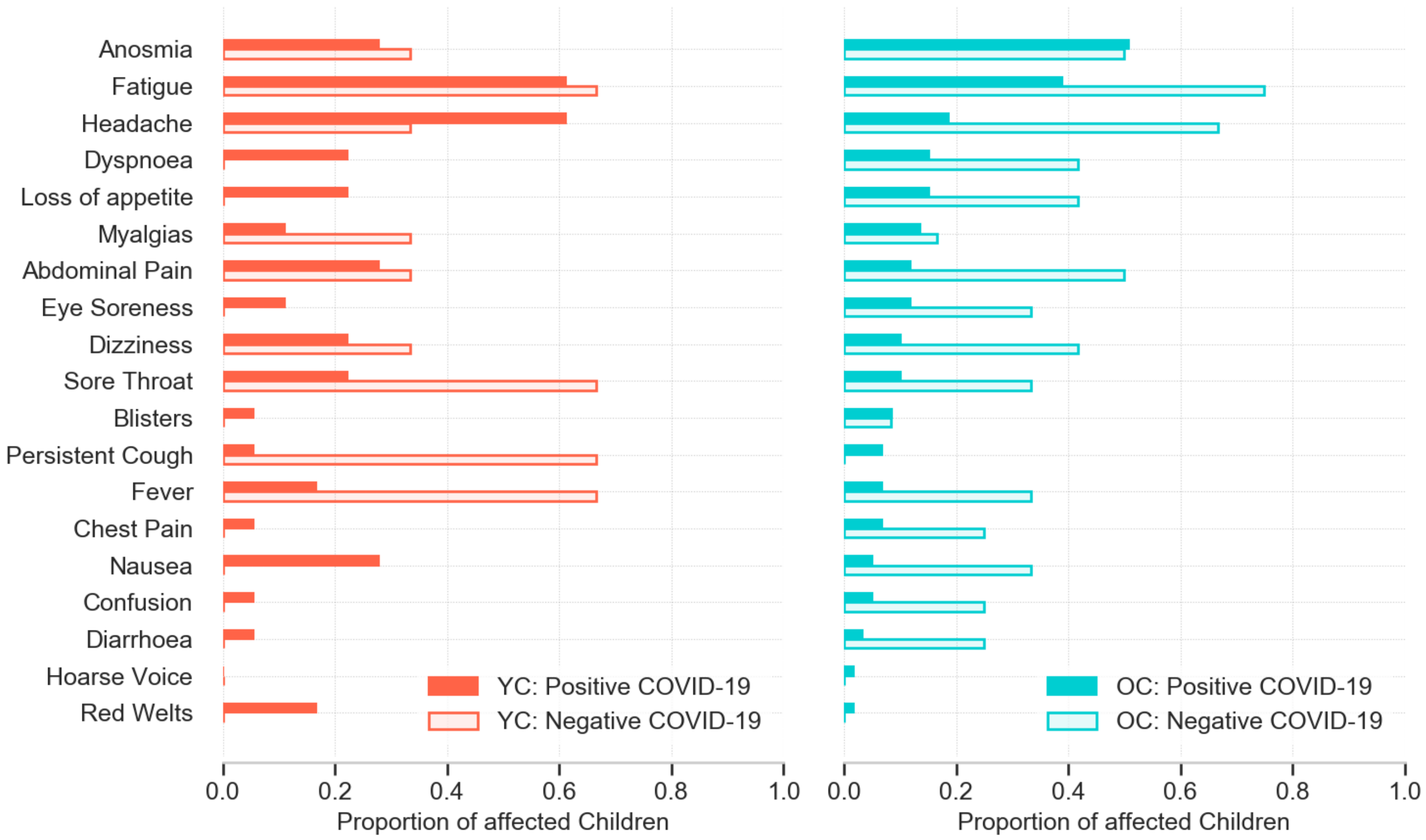

**Supplementary Figure 6. Symptom prevalence over the entire duration of illness in younger children (YC, left panel) and older children (OC, right panel) with illness duration >28 days, comparing children who tested positive (darker bars) or negative (lighter bars) for SARS-CoV-2.**

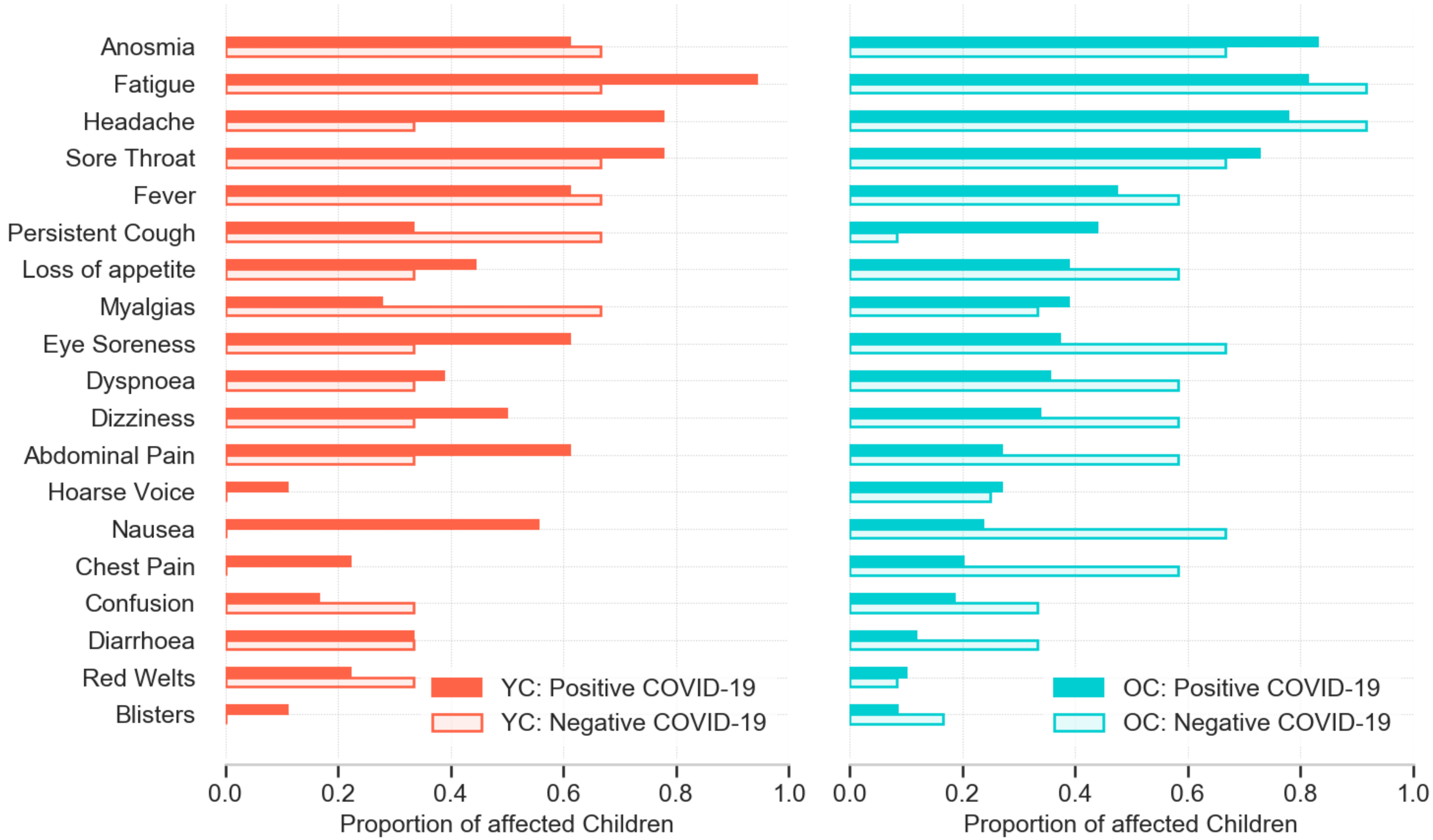
